# Fully-automated sarcopenia assessment in head and neck cancer: development and external validation of a deep learning pipeline

**DOI:** 10.1101/2023.03.01.23286638

**Authors:** Zezhong Ye, Anurag Saraf, Yashwanth Ravipati, Frank Hoebers, Yining Zha, Anna Zapaishchykova, Jirapat Likitlersuang, Roy B. Tishler, Jonathan D. Schoenfeld, Danielle N. Margalit, Robert I. Haddad, Raymond H. Mak, Mohamed Naser, Kareem A. Wahid, Jaakko Sahlsten, Joel Jaskari, Kimmo Kaski, Antti A. Mäkitie, Clifton D. Fuller, Hugo J.W.L. Aerts, Benjamin H. Kann

## Abstract

**Purpose:** Sarcopenia is an established prognostic factor in patients diagnosed with head and neck squamous cell carcinoma (HNSCC). The quantification of sarcopenia assessed by imaging is typically achieved through the skeletal muscle index (SMI), which can be derived from cervical neck skeletal muscle (SM) segmentation and cross-sectional area. However, manual SM segmentation is labor-intensive, prone to inter-observer variability, and impractical for large-scale clinical use. To overcome this challenge, we have developed and externally validated a fully-automated image-based deep learning (DL) platform for cervical vertebral SM segmentation and SMI calculation, and evaluated the relevance of this with survival and toxicity outcomes.

**Materials and Methods:** 899 patients diagnosed as having HNSCC with CT scans from multiple institutes were included, with 335 cases utilized for training, 96 for validation, 48 for internal testing and 393 for external testing. Ground truth single-slice segmentations of SM at the C3 vertebra level were manually generated by experienced radiation oncologists. To develop an efficient method of segmenting the SM, a multi-stage DL pipeline was implemented, consisting of a 2D convolutional neural network (CNN) to select the middle slice of C3 section and a 2D U-Net to segment SM areas. The model performance was evaluated using the Dice Similarity Coefficient (DSC) as the primary metric for the internal test set, and for the external test set the quality of automated segmentation was assessed manually by two experienced radiation oncologists. The L3 skeletal muscle area (SMA) and SMI were then calculated from the C3 cross sectional area (CSA) of the auto-segmented SM. Finally, established SMI cut-offs were used to perform further analyses to assess the correlation with survival and toxicity endpoints in the external institution with univariable and multivariable Cox regression.

**Results:** DSCs for validation set (n = 96) and internal test set (n = 48) were 0.90 (95% CI: 0.90 – 0.91) and 0.90 (95% CI: 0.89 - 0.91), respectively. The predicted CSA is highly correlated with the ground-truth CSA in both validation (r = 0.99, *p* < 0.0001) and test sets (r = 0.96, *p* < 0.0001). In the external test set (n = 377), 96.2% of the SM segmentations were deemed acceptable by consensus expert review. Predicted SMA and SMI values were highly correlated with the ground-truth values, with Pearson r ≥ 0.99 (p < 0.0001) for both the female and male patients in all datasets. Sarcopenia was associated with worse OS (HR 2.05 [95% CI 1.04 - 4.04], p = 0.04) and longer PEG tube duration (median 162 days vs. 134 days, HR 1.51 [95% CI 1.12 - 2.08], p = 0.006 in multivariate analysis.

**Conclusion:** We developed and externally validated a fully-automated platform that strongly correlates with imaging-assessed sarcopenia in patients with H&N cancer that correlates with survival and toxicity outcomes. This study constitutes a significant stride towards the integration of sarcopenia assessment into decision-making for individuals diagnosed with HNSCC.

**SUMMARY STATEMENT:** In this study, we developed and externally validated a deep learning model to investigate the impact of sarcopenia, defined as the loss of skeletal muscle mass, on patients with head and neck squamous cell carcinoma (HNSCC) undergoing radiotherapy. We demonstrated an efficient, fully-automated deep learning pipeline that can accurately segment C3 skeletal muscle area, calculate cross-sectional area, and derive a skeletal muscle index to diagnose sarcopenia from a standard of care CT scan. In multi-institutional data, we found that pre-treatment sarcopenia was associated with significantly reduced overall survival and an increased risk of adverse events. Given the increased vulnerability of patients with HNSCC, the assessment of sarcopenia prior to radiotherapy may aid in informed treatment decision-making and serve as a predictive marker for the necessity of early supportive measures.

## INTRODUCTION

Head and neck squamous cell carcinoma (HNSCC) is the 6^th^ most common cancer worldwide (1), commonly linked to the consumption of alcohol and tobacco and human papillomavirus (HPV) infection (2). The primary treatment approach for early stages is either surgery or radiotherapy, while advanced stages require multimodal therapy, including systemic therapy (3). Although HNSCC can be cured in a portion of locally advanced disease, treatment often results in significant, acute and long-term toxicities (4). Sarcopenia, a skeletal muscle (SM) disorder characterized by decreased muscle function and reduced SM mass, is predominantly found in older adults due to age-related muscle loss and it may also be caused by factors such as malnutrition, inactivity, neurological disorders, and malignant neoplasms (5,6). Progressive sarcopenia is part of cancer cachexia, a multifactorial syndrome that leads to functional decline even with or without loss of fat mass and it cannot be fully reversed by nutritional support (7). Both sarcopenia and cachexia are symptoms and negative prognostic indicators in many forms of malignancy (8). HNSCC patients are particularly susceptible to sarcopenia due to the impact of disease- and treatment-related malnutrition and dysphagia (9,10). Recent studies have established sarcopenia as a negative predictor of overall survival (OS) in HNSCC patients undergoing treatment (10–14).

Computed tomography (CT) is a well-established method for quantifying body composition and it has been extensively employed in clinical research (15). Imaging-assess sarcopenia has typically been performed by of SM at the L3 vertebra (5,16,17). However, routine CT imaging for head and neck cancer (HNC) patients does not extend to the abdomen, which significantly restricts the feasibility of performing L3 measurements using CT images. To overcome this limitation, a series of recent studies proposed a new method of estimating sarcopenia that utilizes SM at the C3 vertebral level and showed a strong correlation with validated standard L3 SM assessment (10,14,18).

Currently, the calculation of skeletal muscle index (SMI) through C3 muscle segmentation relies primarily on manual or semi-automated techniques (10–12,14,18,19), which can be extremely time-consuming and prone to errors as well as interobserver variability. A fully-automated solution with robust segmentation capabilities is necessary to replace the existing manual or semiautomated methods. Over the past years, a multitude of deep learning (DL) models have been created and extensively utilized for the medical image domain (20–23). Although there have been some recent studies that applied DL techniques to determine SM through abdominal CT scans (24–26), very few of them have ventured into utilizing head and neck imaging. Recently, Naser et al. introduced a multi-stage DL approach for segmenting the C3 region using head and neck CT scans, which showed good model performance and its potential for predicting patient survival (27). Despite this, the dataset used was limited in size and lacked external validation (27).

In this study, we developed and externally validated a fully-automated method for segmenting skeletal muscle to calculate the SMI and identify sarcopenia. We evaluated the clinical relevance of these measurements by assessing the prognostic value of baseline quantification of sarcopenia and its influence on survival outcomes and toxicity outcomes in patients during curative therapy. To achieve this, we developed an end-to-end DL pipeline that incorporates 2D Convolutional Neural Networks (CNN) and U-Net models to accurately localize the C3 vertebra and segment the C3 musculature. By automating the process of imaging-assessed sarcopenia determination, this method shows potential to generate fast, consistent, and precise measurements to facilitate evidence-based clinical decision-making for patients diagnosed with HNSCC.

## MATERIALS AND METHODS

### Study design and datasets

This study was conducted in accordance with the Declaration of Helsinki guidelines and following the Mass General Brigham (MGB) Institutional Review Board (IRB) approval. Waiver of consent was obtained from the MGB IRB prior to research initiation. The model development dataset (n = 479) was curated from one public available, de-identified patient cohort from the MD Anderson Cancer Center (MDACC) via The Cancer Imaging Archive (TCIA) (28). Part of the ground truth segmentations for the development dataset (n = 390) was obtained from a public available dataset (29). The remainder of the images in the development dataset (n = 89) were manually reviewed and segmented at the image axial slice- and scan-level by an experienced radiation oncologist (A.S.) with four years of clinical experience using 3D Slicer software (https://slicer.org). For external validation, 1,316 patients undergoing primary radiation therapy for HNSCC from 1996 to 2013 with pretreatment CT simulation scans were retrospectively collected from Brigham and Women’s Hospital (BWH). Of these 1,316 patients, 988 patients were included with complete CT scans and annotated clinical data. Subsequently, 610 patients were excluded due to pre-RT surgery (n = 190), missing body mass index (BMI) information (n = 378), leading to a final set of 420 patients used in the external set (Fig. 1).

**Figure 1.**
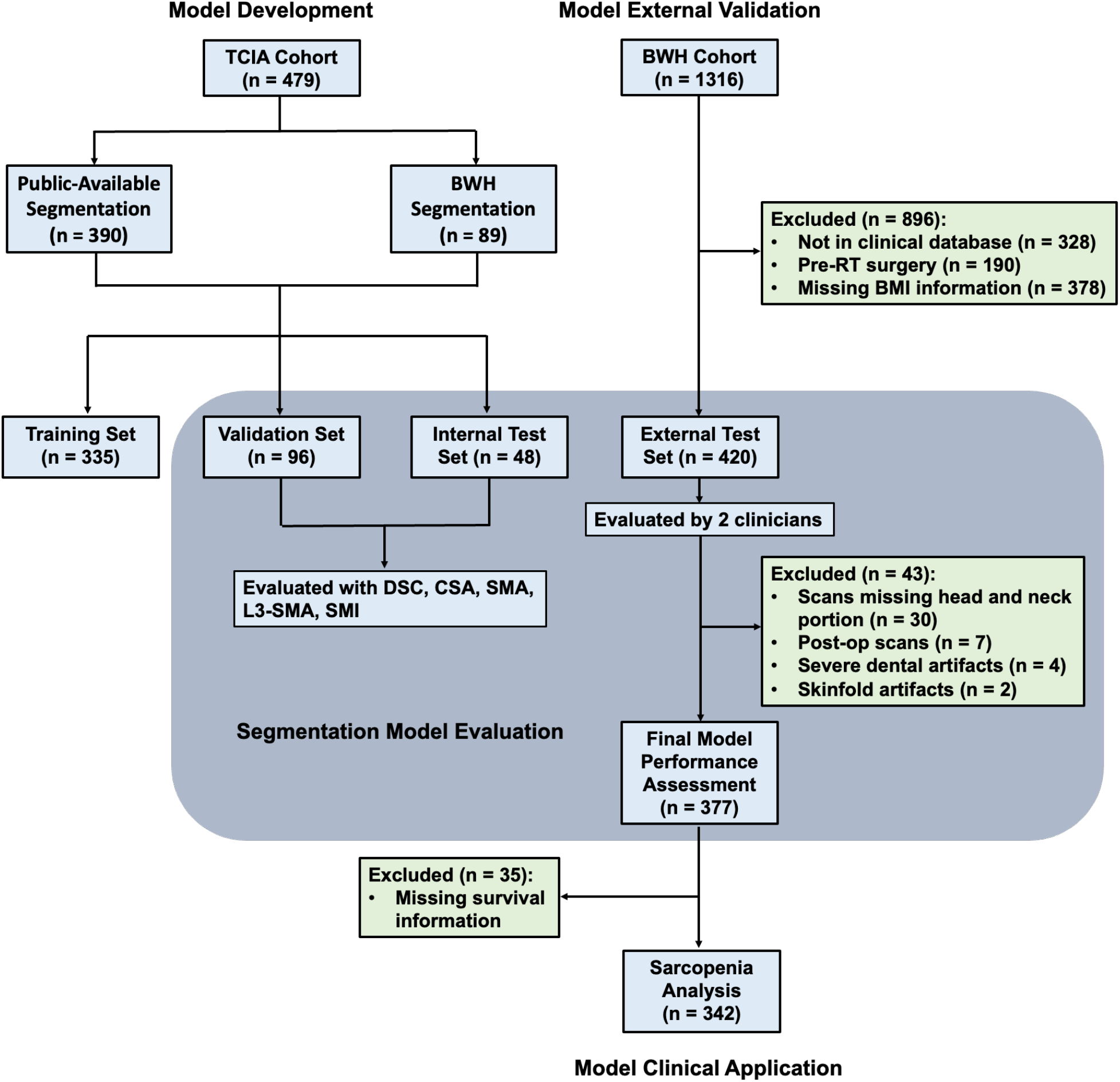
Consort diagrams for training, validation, internal test and external test datasets. TCIA: The Cancer imaging Archive. BWH: Brigham and Women’s Hospital. DSC: dice similarity coefficient. CSA: C3 cross-sectional area; SMA: L3 skeletal muscle cross-sectional area. SMI: skeletal muscle index.

### CT scan characteristics and image acquisition

The CT scans were performed on various CT scanner models from multiple institutions, including GE LightSpeed Plus, GE Discovery ST, Toshiba Aquilion One, Toshiba Aquilion, Philips GeminiGXL 16, Philips Brilliance Big Bore, and TomoTherapy Incorporated Hi-Art. CT scans were diagnostic quality, using 120-140 kVp energy, slice thickness of 1-5 mm, and pixel spacing of 0.3-2.7 mm.

### CT image preprocessing

All CT images were converted from DICOM format to NRRD format via rasterization packages utilizing SimpleITK and plastimatch (https://plastimatch.org) in Python v3.8. For slice selection model and segmentation model, we adopted two different preprocessing strategies. In the slice selection step, CT intensities were first truncated in the range of [−175, 275] Hounsfield units to increase soft tissue contrast and then normalized to the range of [-1, 1] scale. Then the 3-dimensional (3D) images were converted to 2-dimensional (2D) Numpy files with corresponding slice indices as model inputs. In the segmentation step, predicted image slice from slice selection step was extracted from normalized CT images. A standard cropping step was then employed on x-y planes to get rid of excessive empty space for each of the scans. All scans were then resized to 512×512 using linear interpolation via SimpleITK and served as the inputs for the segmentation model.

### Deep learning model implementation

To build an efficient fully-automated pipeline for accurate C3 segmentation, we adopted a two-stage DL approach, consisting of a slice selection step and a segmentation step. Specifically, the slice selection step predicts the C3 image slice from the input 3D CT scan and the segmentation step generates the SM segmentation on the predicted C3 slice. The DenseNet architecture (Fig. S1), known for its impressive classification performance (30), was utilized for train the slice selection model. Similarly, the U-Net architecture (Fig. S2), widely recognized for its effectiveness in biomedical image segmentation tasks (31), was employed for train the semantic segmentation of C3 SM on the chosen C3 image slice. An overview of the architecture for the fully automated segmentation pipeline is provided in Fig. 2. In the slice selection step, the DenseNet regression model performed slice-wise regression predictions on each axial slice of the 3D CT scan independently, followed by post-processing to output the target C3 slice. The model takes input CT slice series and learns to predict a single continuous valued output representing the offset of that slice from the target C3 slice (z-offset). To adapt the model architecture for regression task, the final fully connected layer with softmax activation was replaced with a fully connected layer with a single output unit and a sigmoid activation function to output a number ranging from 0 to 1. The mean absolute error loss between this output and the regression target, C3 slice with 0 z-offset, was then used as the loss function in model training. In the segmentation step, the predicted C3 slice was passed to the U-Net segmentation model to segment the C3 SM for estimating the cross-sectional areas at the C3 level. In the U-Net structure, batch normalization was added to each activation, and the loss function was changed to soft Dice maximization loss in order to deal with class imbalances between the muscle mass and the background.

**Figure 2.**
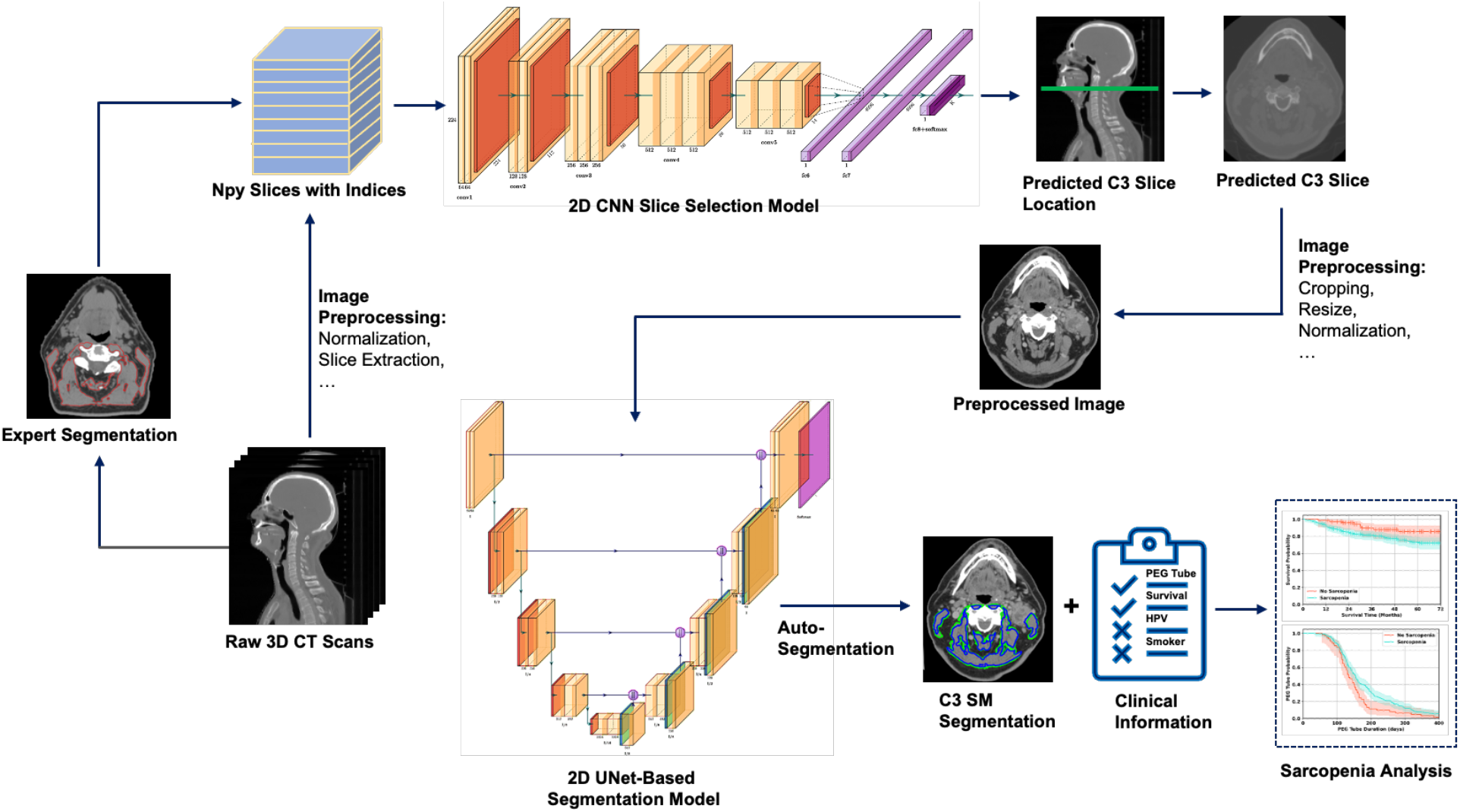
Workflow of the fully-automated deep learning pipeline for accurate C3 segmentation. The 3-dimensional (3D) CT scans were first normalized and converted to 2-dimensional (2D) Numpy files with corresponding slice indices as inputs for slice selection model. The DenseNet regression model performed predictions on each individual axial slice, and then processed the results to determine the target C3 slice. The input to the model is a series of CT slices, and it learns to predict a single continuous value that represents the difference in position (z-offset) of the slice from the target C3 slice. Then the C3 slice was extracted from 3D CT scan and underwent a series of preprocessing steps including cropping, resizing and normalization. Subsequently the preprocessed C3 slice was fed into the U-Net segmentation model to segment the C3 SM and calculate the cross-sectional areas at the C3 level. The L3-SMI was derived to perform a series of predictive analyses.

### Deep learning training and validation

After data preprocessing, the total development dataset (n = 479) was randomly split into training set (n = 335), validation set (n = 96), and test set (n = 48) with a split ratio of 70%:20%:10%. To reduce model overfitting in training, we employed data augmentation strategies including small random translations of up to 0.05 times the image size in both the horizontal and vertical directions, and small rotations of up to 5 degrees in either direction drawn from a uniform distribution. The models were trained for 100 epochs with an initial learning rate of 0.005 that was multiplied by a factor of 0.1 every 25 epochs. To achieve optimal training and validation performance, model hyper-parameters including the number of layers in each dense block of DenseNet, up/down sampling modules, and initial features of U-Net were chosen as recommended in a full body composition study that experimented with a similar architecture by Bridge et al. (32). A batch size of 16 was used for model training and the Adam’s optimizer was used to minimize the loss functions during the training of both models. All models were trained from scratch using TensorFlow v2.8 in Python. The performance of the automated pipeline was evaluated by the placement of the C3 selected slice and the Dice Similarity Coefficient (DSC) of the auto segmentation over ground truth on the validation set. The reliability of the auto segmentations for its use in the sarcopenia determination is evaluated by the Intra Class Correlation (ICC) coefficient of cross-sectional area measurement.

### Model external validation on BWH cohorts

To determine if the model could generalize to patients from outside institutions, we used HNSCC CT scans (n = 420) from BWH dataset for external validation of the model. To evaluate the model C3 segmentation performance, two experienced board-certified head and neck radiation oncologists (B.H.K., 10 years of experience and F.H., 21 years of experience) individually reviewed and evaluated the segmentations with Likert scales of 0 to 3: 0 = wrong slice, 1 = unacceptable (expected segmentation variation of β 5% versus ground truth), 2 = acceptable, minor revisions needed (expected segmentation variation of <5% versus ground truth), 3 = acceptable, no revisions needed. The inter-rater reliability test was performed to measure the agreement.

### Definition of sarcopenia

As proposed by Swartz et al. (18) and van Rijn-Dekker et al. (10), the SMA at the L3 lumbar level was calculated based on Equation 1 and then the SMI was calculated from Equation 2.

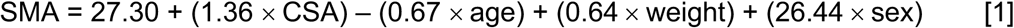

Here SMA is the cross-sectional area in cm^2^ at the L3 lumbar level. CSA is the cross-sectional area in cm^2^ at the C3 cervical level. Age is the patient’s age in years. Weight is the patient’s weight in kg. Sex is equal to 1 if the patient is female and is equal to 2 if the patient is male.

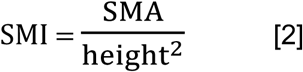

Here SMI_L3_ is SMI at the L3 lumber level. SMA is the SMA in cm^2^ at the L3 lumbar level. Height is the patient’s height in meters.

SMI thresholds of 52.4 cm^2^/m^2^ for males and 38.5 cm^2^/m^2^ for females were adopted for SMI to classify patients into sarcopenia group and non-sarcopenia group, as previously established by Prado et al. (33).

### Statistical analysis

The Dice similarity coefficient scores (DSC) were calculated and served as the primary endpoint to assess the similarity between the model predicted C3 segmented area with the ground truth segmentation. Precision and recall scores were also calculated to provide supplementary information on the model performance. Additionally, Intraclass Correlation Coefficient (ICC) scores were generated to access the model performance. To examine the correlation between the cross-sectional areas, we utilized the Pearson correlation coefficient. To determine if these areas were significantly different, we performed a two-sided Wilcoxon signed-rank test. The Pearson’s Chi-square test was performed to test the statistically significant differences among the training, validation and internal test and external test datasets. A p-value less than 0.05 was considered statistically significant. The inter-rater reliability test was used to measure the agreement between the ratings by two clinicians on the acceptability of SM segmentations for the external validation set. To address the data imbalance of the acceptability scores, we adopted the Agreement Coefficient 1 (AC_1_) introduced by Gwet (34). The 95% confidence intervals were calculated based on 10,000 bootstrapped iterations. All the above statistical metrics and curves were generated using the Scikit-learn and SciPy packages in Python v3.8. The predictive effect of sarcopenia on toxicity endpoints was evaluated using univariate logistic regression analyses. The sarcopenia associations with overall survival (OS) and PEG tube duration were assessed on the external dataset with Cox Proportional Hazard (CoxPH) regression analysis. The overall survival and PEG tube duration was visualized using Kaplan Meier curves with Lifelines package in Python v3.8. All the CoxPH and logistic regression analyses were conducted with Stata, Version 17.0 (College Station, TX, USA).

## RESULTS

### Patient characteristics

The total patient cohort consisted of 899 HNSCC patients from two institutions, with 479 patients in the development set from MD Anderson Cancer Center (MDACC) and 420 patients in the external test set from Brigham and Women’s Hospital (BWH) (Table 1). The age of the patients ranged from 24 to 90 years old, with a median age of 58 years. Most of the patients were male (83.9%, n = 754), as typically found in HNSCC. The primary cancer site was most commonly the oropharynx (84.5%, n = 760), followed by the larynx, hypopharynx, and nasopharynx. Most of the patients had stage IV cancer (73.9%, n = 664) according to the American Joint Committee on Cancer (AJCC) 7th edition staging system, followed by stage III cancer (16.1%, n = 145). The human papillomavirus (HPV)/p16 status was positive for 48.3% (n = 434) of patients, negative for 9.6% (n = 86) of patients, and unspecified for 42.2% (n = 379) of patients.

**Table 1.** Head and neck cancer patient characteristics.

| Patient Cohort<br>(n = 899) | Training<br>(MDACC, n = 335) | Validation<br>(MDACC, n = 96) | Internal Test<br>(MDACC, n = 48) | External Test<br>(BWH, n = 420) | p-value |
| --- | --- | --- | --- | --- | --- |
| **Age** |  |  |  |  | 0.37<sup>#</sup> |
| median (range) | 57 (24 – 83) | 58 (29 – 81) | 59.5 (41 – 87) | 59 (24-87) |  |
| **Sex n (%)** |  |  |  |  |  |
| Female | 41 (12.2%) | 16 (16.7%) | 8 (16.7%) | 75 (17.9%) | 0.32<sup>+</sup> |
| Male | 291 (86.9%) | 80 (83.3%) | 40 (83.3%) | 344 (81.9%) |  |
| Unspecified | 3 (0.9%) | 0 (0.0%) | 0 (0.0%) | 1 (0.2%) |  |
| **Smoking Status** |  |  |  |  | < 0.001<sup>+</sup> |
| Current | 68 (20.3%) | 19 (19.8%) | 11 (22.9%) | 55 (13.1%) |  |
| Former | 95 (28.4%) | 40 (41.7%) | 17 (35.4%) | 217 (51.7%) |  |
| Never | 103 (30.7%) | 33 (34.4%) | 17 (35.4%) | 145 (34.5%) |  |
| Unspecified | 67 (20.0%) | 4 (4.2%) | 3 (6.2%) | 3 (0.7%) |  |
| **Primary Cancer Site n (%)** |  |  |  |  | < 0.001<sup>+</sup> |
| Oropharynx | 309 (92.2%) | 92 (95.8%) | 45 (93.8%) | 314 (74.8%) |  |
| Nasopharynx | 1 (0.3%) | 0 (0.0%) | 0 (0.0%) | 0 (0.0%) |  |
| Larynx/Hypopharynx | 2 (0.6%) | 0 (0.0%) | 0 (0.0%) | 76 (18.1%) |  |
| Oral cavity | 2 (0.6%) | 0 (0.0%) | 0 (0.0%) | 0 (0.0%) |  |
| Unknown/Other | 19 (5.7%) | 4 (4.2%) | 3 (6.2%) | 30 (7.1%) |  |
| **AJCC Stage (7<sup>th</sup> ed) n (%)** |  |  |  |  | 0.001<sup>+</sup> |
| I | 3 (0.9%) | 1 (1.0%) | 0 (0.0%) | 12 (2.9%) |  |
| II | 11 (3.3%) | 3 (3.1%) | 2 (4.2%) | 34 (8.1%) |  |
| III | 42 (12.5%) | 16 (16.7%) | 12 (25.0%) | 75 (17.9%) |  |
| IV | 266 (79.4%) | 72 (75.0%) | 31 (64.6%) | 295 (70.2%) |  |
| Unspecified | 13 (3.9%) | 4 (4.2%) | 3 (6.2%) | 4 (1.0%) |  |
| **HPV/p16 Status\* n (%)** |  |  |  |  | < 0.001<sup>+</sup> |
| Negative | 19 (5.7%) | 9 (9.4%) | 9 (18.8%) | 49 (11.7%) |  |
| Positive | 98 (29.3%) | 76 (79.2%) | 36 (75.0%) | 224 (53.3%) |  |
| Unspecified | 218 (65.1%) | 11 (11.5%) | 3 (6.2%) | 147 (35.0%) |  |
| **T-Stage (7<sup>th</sup> ed) n (%)** |  |  |  |  | 0.24<sup>+</sup> |
| T0 | 2 (0.6%) | 0 (0.0%) | 0 (0.0%) | 0 (0.0%) |  |
| T1 | 56 (16.7%) | 20 (20.8%) | 11 (22.9%) | 89 (21.2%) |  |
| T2 | 143 (42.7%) | 35 (36.5%) | 17 (35.4%) | 160 (38.1%) |  |
| T3 | 77 (23.0%) | 22 (22.9%) | 10 (20.8%) | 109 (26.0%) |  |
| T4 | 44 (13.1%) | 15 (15.6%) | 7 (14.6%) | 57 (13.6%) |  |
| Unspecified | 13 (3.9%) | 4 (4.2%) | 3 (6.2%) | 5 (1.2%) |  |
| **N-Stage (7<sup>th</sup> ed)**<br>**n (%)** |  |  |  |  | < 0.001<sup>+</sup> |
| N0 | 33 (9.9%) | 9 (9.4%) | 5 (10.4%) | 85 (20.2%) |  |
| N1 | 31 (9.3%) | 13 (13.5%) | 9 (18.8%) | 52 (12.4%) |  |
| N2 | 246 (74.3%) | 68 (70.8%) | 31 (64.6%) | 247 (58.8%) |  |
| N3 | 12 (3.6%) | 2 (2.1%) | 0 (0.0%) | 31 (7.4%) |  |
| Unspecified | 13 (3.9%) | 4 (4.2%) | 3 (6.2%) | 5 (1.2%) |  |

*\*Patients with non-oro-pharyngeal carcinoma who did not undergo HPV- or p16-testing were coded as negative, given the very low incidence of HPV/p16 positive tumors in these disease sites. For numerical data as in age, the Kruskal-Wallis rank sum test (#) was performed to test the statistical significances among age medians. For categorical data, the Fisher's Exact test (+) was performed to test the statistically significant differences among train, validation and internal test and external test datasets. AJCC: American Joint Committee on Cancer. HPV: Human papillomavirus.*

### Slice selection and auto-segmentation model performance

Evaluation of model slice selection revealed that the difference between the predicted mid-C3 slice and the ground truth slice was minimal, as demonstrated by the histogram analyses of the difference between the locations of the predicted C3 slice and the ground truth slice (Fig. 3A). The mean difference (Δh) between the locations of the predicted C3 slice and the ground truth slice was 0.11 ± 1.13 mm and 0.07 ± 1.08 mm for validation and internal test sets, respectively (Fig. 3A). The DSC values obtained for the validation set and internal test set predicted segmentations as compared to ground truth were found to be 0.90 (95% CI 0.90 - 0.91) and 0.90 (95% CI 0.89 - 0.91), respectively (Fig. 3C). Additionally, the precision (validation: 0.97 [95% CI 0.96 - 0.97]; test: 0.97 [95% CI 0.95 – 0.97]), recall (validation: 0.84 [95% CI 0.84 - 0.85], test: 0.85 [95% CI 0.83 - 0.85]) and ICC scores (validation: 0.99 [95% CI 0.98 - 0.99], test: 0.96 [95% CI 0.94 - 0.98]) as summarized in Fig. 3D all showed excellent model performance in predicting C3 segmentations. C3 CSAs derived from predicted segmentations showed near perfect correlations with the ground truth calculated CSAs (validation: r = 0.99, *p* < 0.0001, test: r = 0.96, *p* < 0.0001, Fig 3B-C). Representative examples of C3 section slices on sagittal CT images and ground truth segmentations on axial images with performance metrics are shown in Fig. 4.

**Figure 3.**
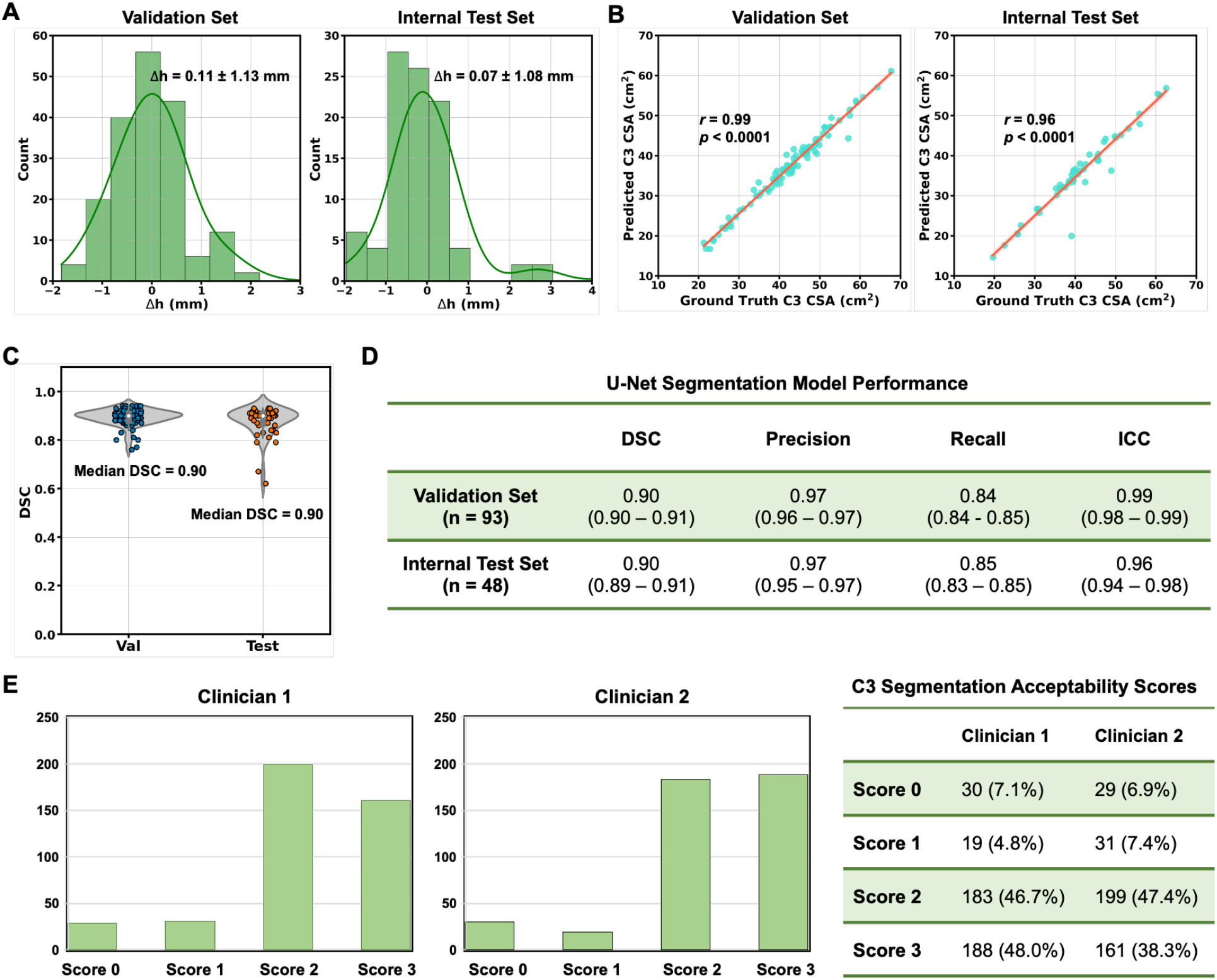
The performance of the CNN slice selection model and U-Net segmentation model was evaluated for the segmentation of the C3 vertebra section. (A) Histogram shows the difference (Δh) between the location of the model-predicted C3 section slice and the location of the ground-truth manually segmented CT slice for validation set (mean Δh = 0.11 ± 1.13 mm) and internal test set (mean Δh = 0.07 ± 1.08 mm). (B) Scatter plot depicts the C3 skeletal muscle cross-sectional area (CSA, cm^2^), with the ground-truth manual segmentation on the x-axis and the calculated CSA (cm^2^) using predicted segmentations on the y-axis for validation and internal test sets. (C) DSC distributions were shown for validation and internal test sets. (D) DSC, precision, recall and intra-class correlation coefficient values were summarized to show the model performance. (E) C3 segmentations predicted by model were individually reviewed by two experienced board-certified radiation oncologists with Likert scales of 0 - 3: 0 - wrong slice, 1 - unacceptable (expected segmentation variation of ≥ 5% versus ground truth), 2 - acceptable, minor revisions needed (expected segmentation variation of < 5% versus ground truth), 3 - acceptable, no revisions needed. IQR: inter quantile range. DSC: Dice similarity coefficient.

**Figure 4.**
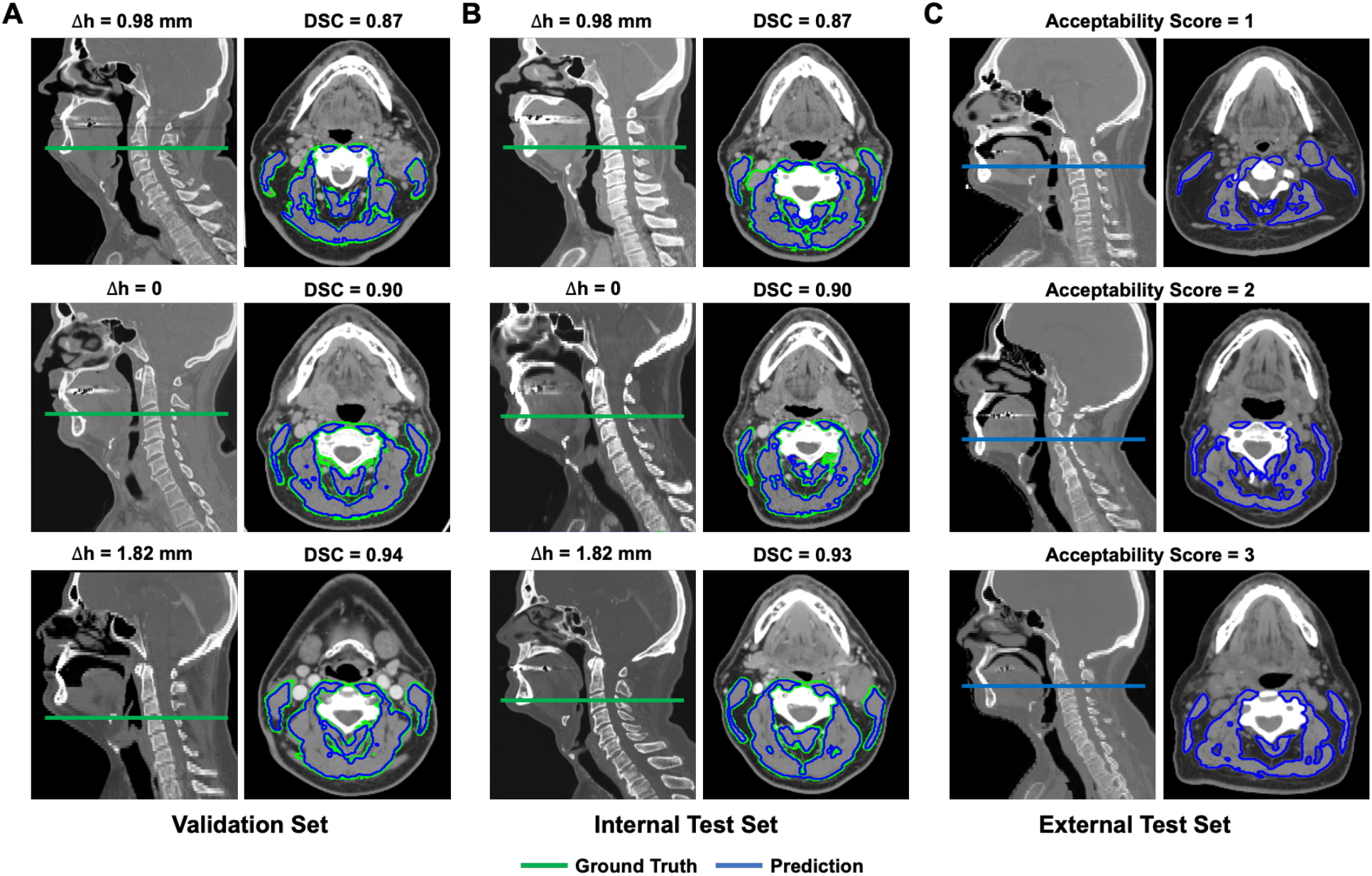
Representative cases with ground truth slices of C3 sections on sagittal CT images (green plane) and ground truth segmentations (green contours) on axial images were shown for validation (A) and internal test (B) sets. Predicted segmentations (blue contours) with varying DSC values (> median DSC, = median DSC, < median DSC) were also overlaid on axial CT images as compared to show their similarities to ground truth segmentations for validation (A) and internal test (B) sets. Corresponding distance (Δh) between the predicted C3 section slice and ground-truth slice and DSC values were annotated for each case in validation (A) and internal test sets (B). Model predicted C3 slices on sagittal CT images (blue planes) and segmentation on axial CT images (blue contours) were also shown for the external test set (C). Acceptability scores from expert clinicians’ review were annotated for corresponding cases in the external test set (C). DSC: Dice similarity coefficient.

### Skeletal muscle index measurement comparisons

We calculated and compared the SMA and SMI values between model predictions and ground truth for both the validation set (Fig. 5A) and internal test set (Fig. 5B). Accurate model SM segmentations led to predicted SMA and SMI values were highly correlated with the ground-truth values, with Pearson r ≥ 0.99 (p < 0.0001) for both female and male patients in all datasets (Fig. 5).

**Figure 5.**
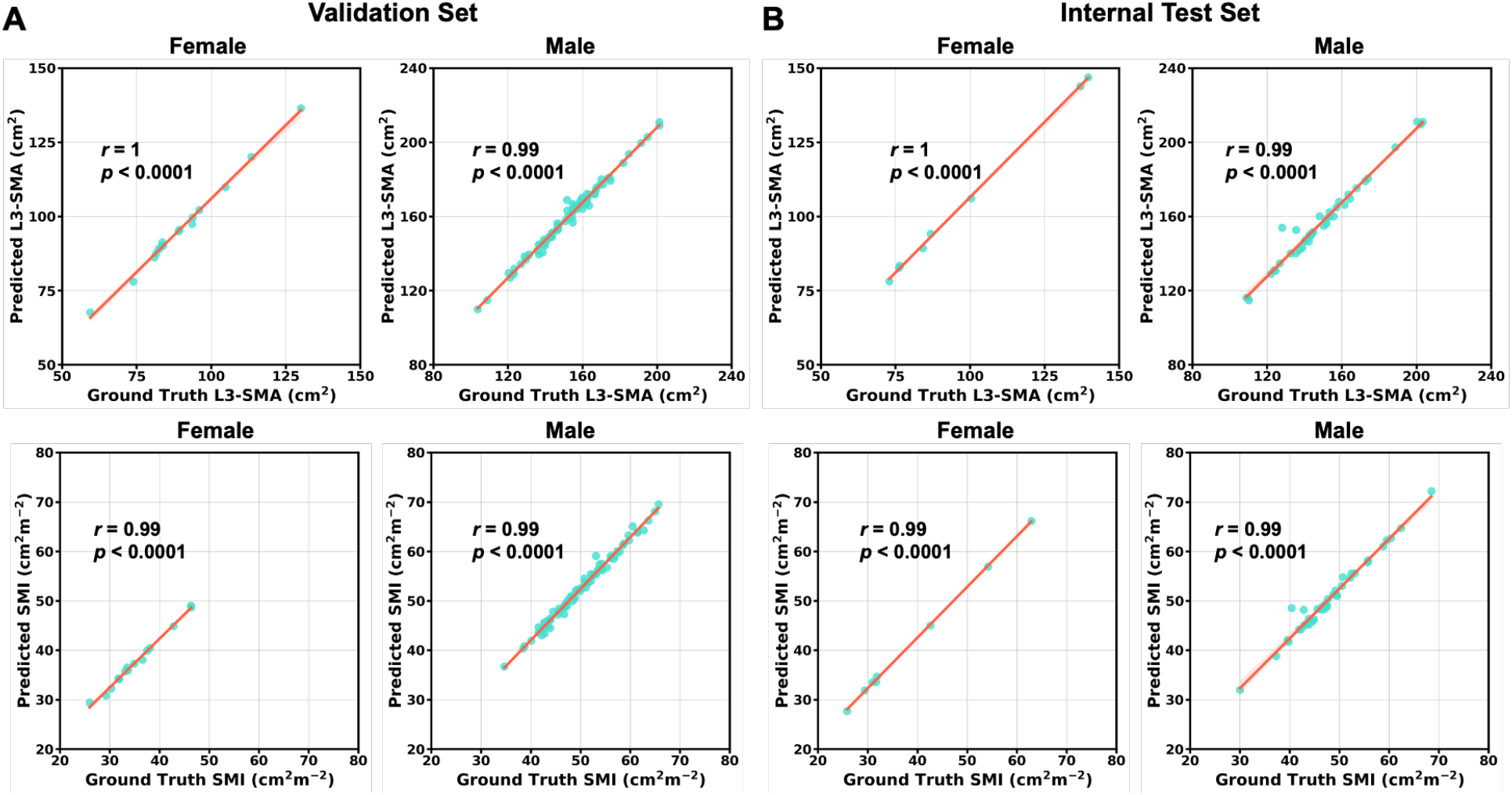
Scatter plots of the lumbar level 3 skeletal muscle cross-sectional area (SMA) and skeletal muscle index (SMI) values determined for validation set (A) and internal test set (B) patients (stratified by sex) using the ground-truth manual segmentation (x-axis) and model predicted segmentations (y-axis). Pearson’s correlations showed all model-predicted values and ground truth values were significantly correlated (p < 0.0001).

### Evaluation on external test set

Representative cases with C3 slice predictions and segmentations for acceptability scores 1, 2, 3 are shown in Fig. 4C for the external test set. The review scores were summarized in Fig. 3E, with 183 (46.7%) and 188 (48.9%) cases deemed acceptable with minor or no changes needed, respectively for reviewer 1 and with 199 (47.4%) and 161 (38.3%) cases deemed acceptable with minor or no changes needed, respectively for reviewer 2. We qualitatively re-reviewed all unacceptable cases and identified 42 (10%) problematic scans, including those without head and neck portion (n = 30), post-operative (post neck-dissection) scans (n = 7), severe dental artifact (n = 4), skinfold artifact (n = 2). After exclusion of the faulty scans, we had a final set of 377 patients and high inter-rater agreement (AC_1_ score = 0.94). We further investigated the unacceptable segmentations that were given by either one of the reviewers. We identified 23 cases (6.1%), with 11 cases (2.9%) from reviewer 1 and 18 cases (4.8%) from reviewer 2. Failure modes are summarized in Table S1, and included 9 cases (39.1%) with sternocleidomastoid (SCM) muscle missing (Fig. S3A); 6 cases (26.1%) with lymph node included (Fig. S3B); 3 cases (13.0%) with posterior neck muscles missing (Fig. S3C); 3 cases (13%) with anterior deep muscle missing (Fig. S3D); 1 case (4.4%) with submental muscle included (Fig. S3E); and 1 case (4.4%) with other muscle included (Fig. S3F). Given high overall acceptability, we moved forward with SMI calculation and designation of sarcopenia for the external test set.

### Univariate and multivariate analyses for Sarcopenia

A total of 342 patients with complete survival and toxicity information from the external test set were further included for sarcopenia predictive analysis (Table 2). The median follow-up for all patients was 43 months (range 1 month – 170 month). The overall survival at 5 years was 80.7%. There were 261 (76.3%) sarcopenic patients and 81 (23.7%) non-sarcopenic patients in the dataset. Median age was 59 (range 24 - 87), most were male (83%), smoking history <10 pack-years (py) (51%), Adult Comorbidity Evaluation 27 (ACE-27) score 0 (39%) or 1 (38%), non-oropharynx primary (73%), and American Joint Committee on Cancer (AJCC) 7th edition stage III (16%), IVa (65%), or IVb (8%).

**Table 2.**
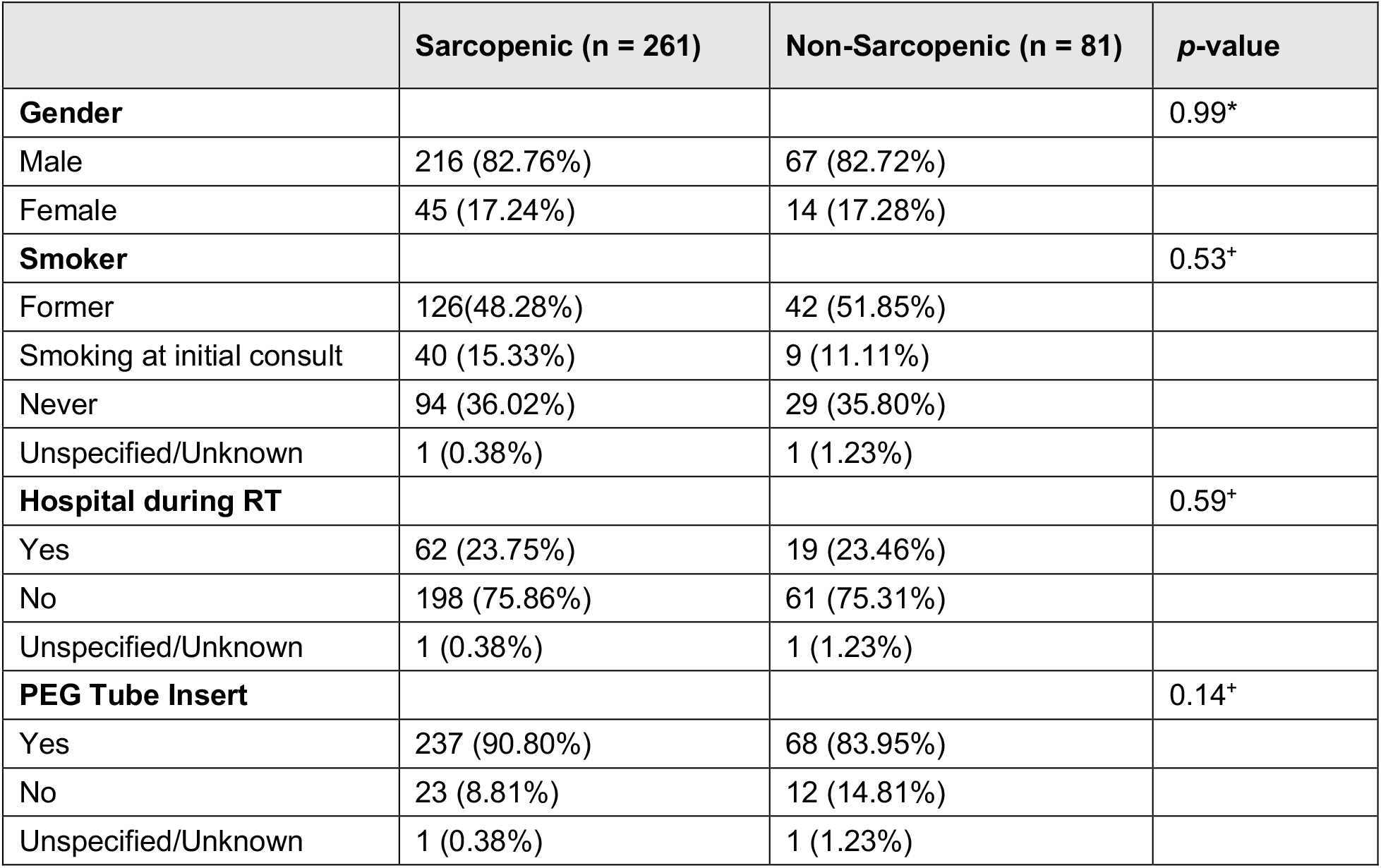
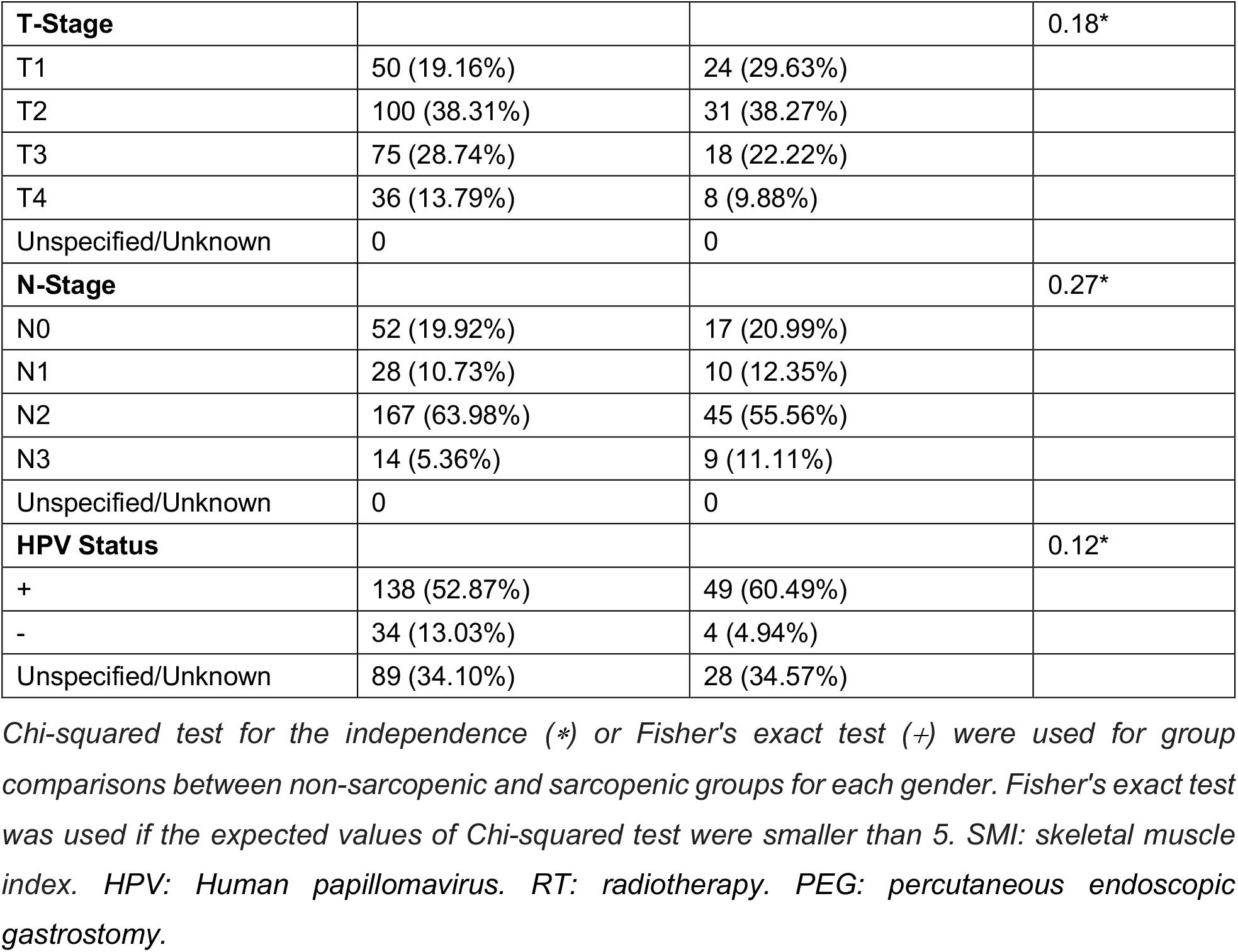
Patient characteristics for Non-Sarcopenic and Sarcopenic groups.

### Sarcopenia and survival outcomes

Five-year survival was 84.4% in patients without sarcopenia vs. 73.1% in patients with sarcopenia (HR: 2.21 [95% CI 1.08 - 4.12], *p* = 0.03) (Fig. 6A). On multivariate analysis, variables associated with worse OS were sarcopenia (HR 2.05 [95% CI 1.04 - 4.04], *p* = 0.04), ACE-27 score 2+ (HR 2.24 [95% CI 1.39 - 3.62], *p* = 0.001), non-oropharynx diagnosis (HR 3.92 [95% CI 2.45 - 6.25], *p* < 0.001), and T3-4 stage (HR 2.36 [95% CI 1.47 - 3.77], *p* < 0.001), but not age > 65 (*p* = 0.79), smoking history >10py (*p* = 0.75), N2-3 (*p* = 0.60), or stage 3 - 4 (p = 0.41) (Table 3).

**Table 3.**
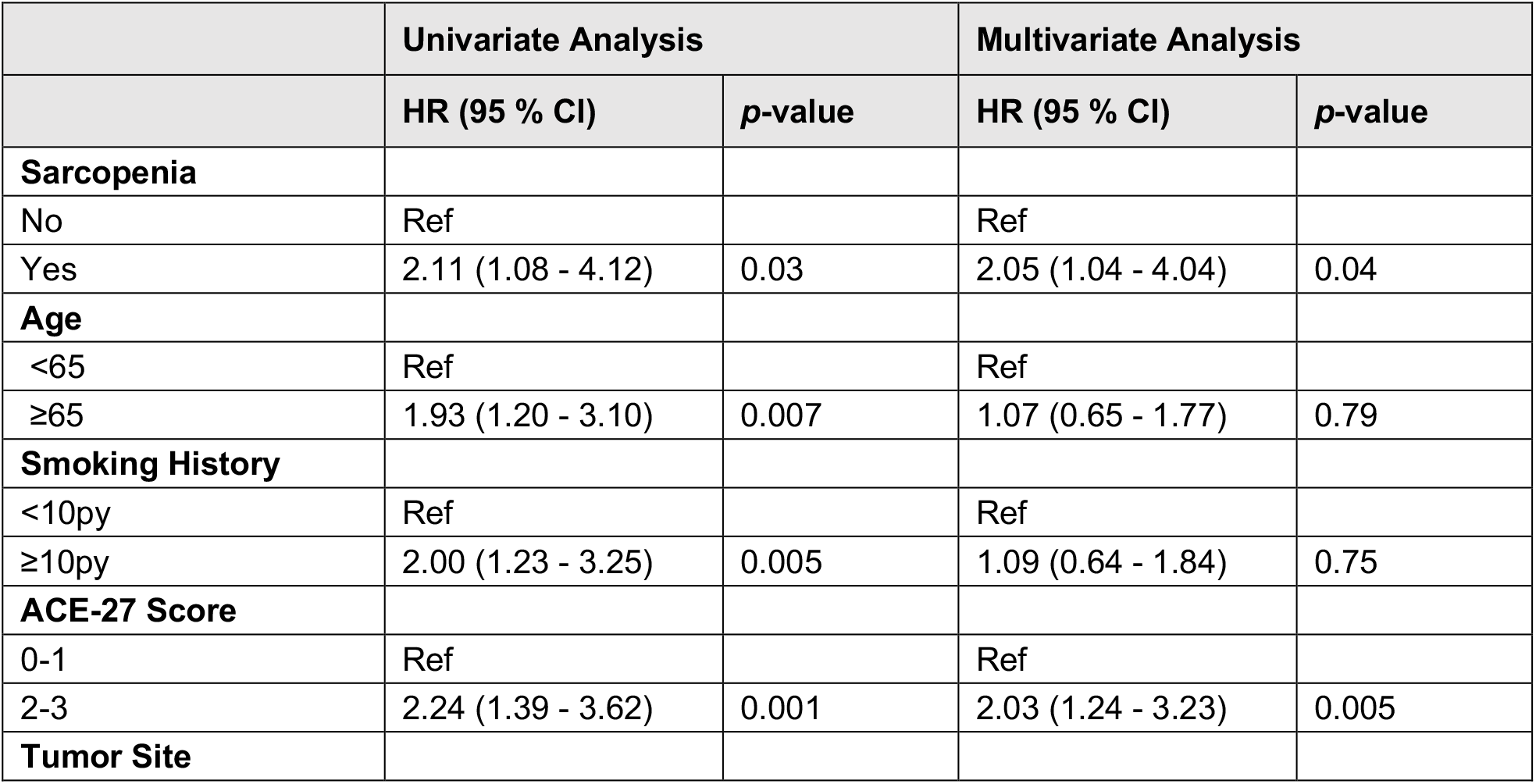
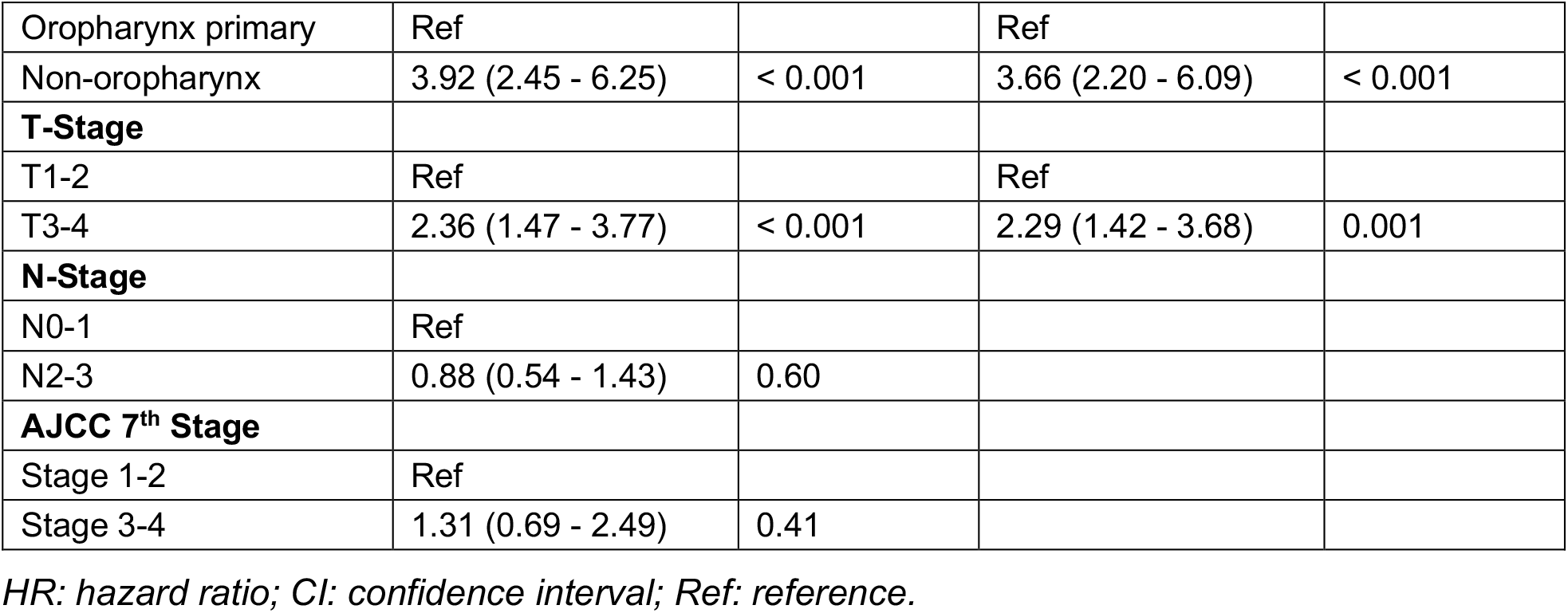
Univariate and multivariate analyses for overall survival.

|  | Univariate Analysis |  | Multivariate Analysis |  |
| --- | --- | --- | --- | --- |
|  | HR (95 % CI) | p-value | HR (95 % CI) | p-value |
| **Sarcopenia** |  |  |  |  |
| No | Ref |  | Ref |  |
| Yes | 2.11 (1.08 - 4.12) | 0.03 | 2.05 (1.04 - 4.04) | 0.04 |
| **Age** |  |  |  |  |
| <65 | Ref |  | Ref |  |
| ≥65 | 1.93 (1.20 - 3.10) | 0.007 | 1.07 (0.65 - 1.77) | 0.79 |
| **Smoking History** |  |  |  |  |
| <10py | Ref |  | Ref |  |
| ≥10py | 2.00 (1.23 - 3.25) | 0.005 | 1.09 (0.64 - 1.84) | 0.75 |
| **ACE-27 Score** |  |  |  |  |
| 0-1 | Ref |  | Ref |  |
| 2-3 | 2.24 (1.39 - 3.62) | 0.001 | 2.03 (1.24 - 3.23) | 0.005 |
| **Tumor Site** |  |  |  |  |
| Oropharynx primary | Ref |  | Ref |  |
| Non-oropharynx | 3.92 (2.45 - 6.25) | < 0.001 | 3.66 (2.20 - 6.09) | < 0.001 |
| **T-Stage** |  |  |  |  |
| T1-2 | Ref |  | Ref |  |
| T3-4 | 2.36 (1.47 - 3.77) | < 0.001 | 2.29 (1.42 - 3.68) | 0.001 |
| **N-Stage** |  |  |  |  |
| N0-1 | Ref |  |  |  |
| N2-3 | 0.88 (0.54 - 1.43) | 0.60 |  |  |
| **AJCC 7<sup>th</sup> Stage** |  |  |  |  |
| Stage 1-2 | Ref |  |  |  |
| Stage 3-4 | 1.31 (0.69 - 2.49) | 0.41 |  |  |

HR: hazard ratio; CI: confidence interval; Ref: reference.

**Figure 6.**
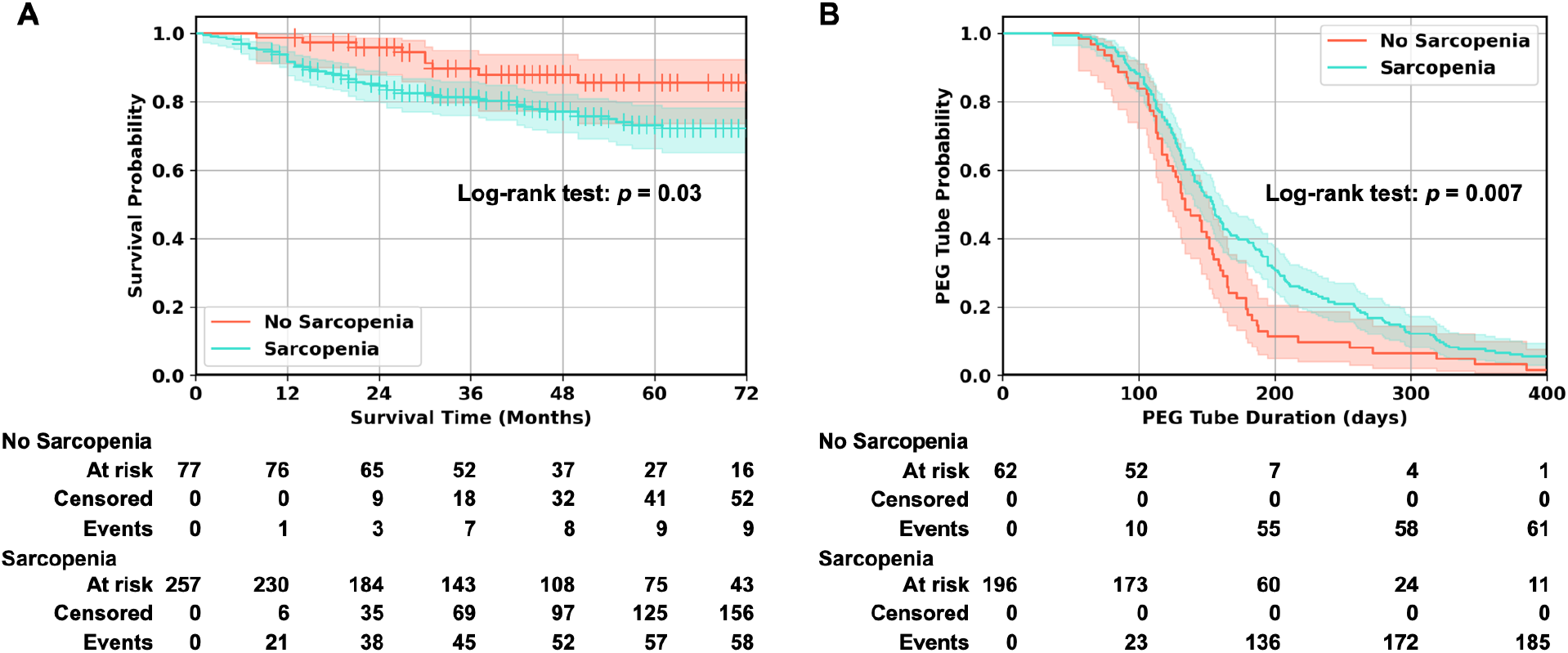
Kaplan-Meier curves show significant differences in both the overall survival time (Log-rank test p = 0.03) and the PEG tube duration (Log-rank test p = 0.007) between sarcopenia patients and no sarcopenia patients.

### Sarcopenia and toxicity outcomes

Sarcopenia was associated with longer percutaneous endoscopic gastrostomy (PEG) tube duration (median 162 days vs. 134 days, HR 1.67 [95% CI 1.23 - 2.22], *p* = 0.001) (Table 4, Fig. 6B). On multivariate analysis, variables associated with longer PEG tube duration were sarcopenia (HR 0.66 [95% CI 0.48 – 0.89], *p* = 0.003), ACE-27 score (HR 0.72 [95% CI 0.53 – 0.97], *p* = 0.03) and non-oropharynx primary site (HR 0.80 [95% CI 0.56 - 1.14], *p* = 0.03) (Table 4). Sarcopenia was not associated with insertion of PEG tube at diagnosis (*p* = 0.12) but was associated with higher risk of having PEG tube at last follow-up (odds ratio (OR) 2.25 [95% CI 1.02 - 4.99], *p* = 0.05) (Table 5). Sarcopenia was not significantly associated with higher risk of hospitalization < 3 months after RT (Table 5; OR 2.18 [95% CI 0.82 - 5.79], *p* = 0.12). Sarcopenia was not significantly associated with risk of osteoradionecrosis (*p* = 0.39), post-RT stricture (*p* = 0.24), or treatment-complication requiring surgery (*p* = 0.50) (Table 5).

**Table 4.**
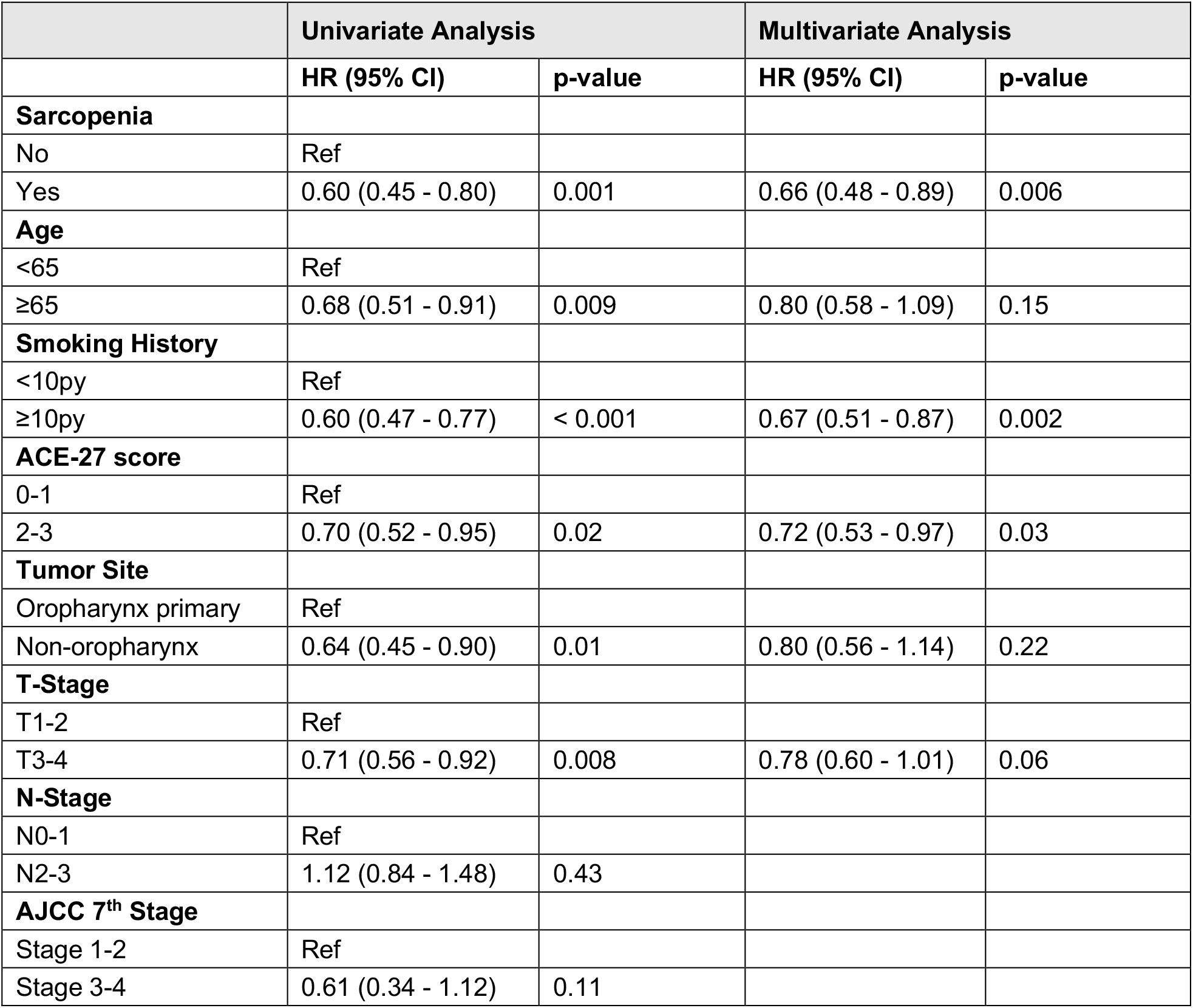
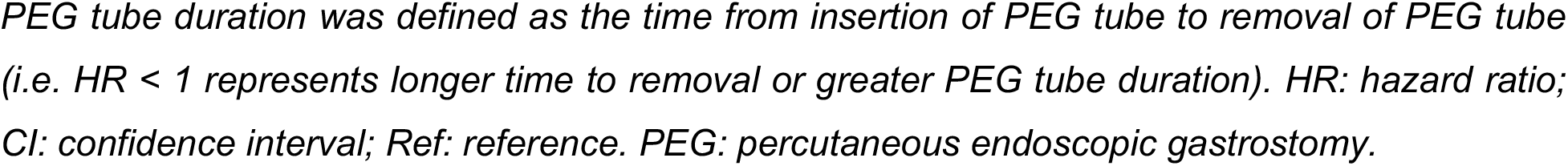
Univariate and multivariate analyses for PEG tube duration.

**Table 5.** Univariate analysis for the association of sarcopenia with various toxicity endpoints.

| Toxicity | No Sarcopenia | Sarcopenia | OR (95% CI) | p-value |
| --- | --- | --- | --- | --- |
| PEG inserted at diagnosis | 84% | 91% | 1.81 (0.86 - 3.84) | 0.12 |
| PEG removal | 77% | 75% | 0.46 (0.19 - 1.14) | 0.09 |
| PEG at last follow up | 10% | 18% | 2.25 (1.02 - 4.99) | 0.05 |
| Hospitalization during RT | 23% | 24% | 1.01 (0.56 - 1.81) | 0.97 |
| Hospitalization <3months after RT | 6% | 13% | 2.18 (0.82 - 5.79) | 0.12 |
| Osteoradionecrosis | 2% | 1% | 0.45 (0.07 - 2.77) | 0.39 |
| Post-RT stricture | 7% | 12% | 1.73 (0.70 - 4.30) | 0.24 |
| Complication from treatment requiring surgery | 7% | 10% | 1.38 (0.55 - 3.47) | 0.50 |

OR: odds ratio; RT: radiotherapy; PEG: percutaneous endoscopic gastrostomy.

## DISCUSSION

In this study, we successfully developed and validated an end-to-end DL pipeline that utilizes head and neck CT images for efficient and accurate segmentation of cervical vertebral SM, calculation of SMI, and diagnosis of imaging-based sarcopenia in HNSCC patients. Our tool was applied to a large external validation cohort, where we found that imaging-based sarcopenia was associated with poorer OS and longer PEG tube duration. This externally-validated DL pipeline offers significant promise for clinical translation as a fast and fully-automated prognostic tool for the HNSCC patients in routine clinical practice. This is the first end-to-end DL pipeline for determining sarcopenia that uses head and neck CT images and has been externally validated with a substantial patient population.

We followed a two-step process, which is similar to a recent study conducted by Naser et al. (27), to segment the C3 SM using two separate DL models. However, our methods differ significantly. Naser et al. used a 3D ResUNet model to segment the C3 vertebra section initially. They then automatically selected the middle slice of the section and applied a 2D ResUNet model to segment the SM on the selected slice. In contrast, we used a 2D DenseNet-based regression model to automatically select the C3 SM slice (slice selection model). We then used a 2D U-Net model to segment the selected slice (segmentation model). We achieved excellent model performance for both slice selection and segmentation models in the validation and internal test sets. In a large external test set, 96.2% of the SM segmentations were also deemed acceptable by expert consensus review. Compared to 3D CNN models, 2D CNN models are generally much easier to train and implement, making our pipeline fast and efficient for the C3 SM segmentation for sarcopenia analysis. Typically, it took an experienced board-certified radiation oncologist 5 - 10 minutes to identify and segment a C3 SM. In contrast, our end-to-end DL pipeline was much faster and only required 0.15 seconds to segment the C3 SM. This is considerably quicker than a human expert.

Sarcopenia has been found to be a significant prognostic factor for decreased overall survival in patients with varying types of cancers (5,6,33). For head and neck cancer, these findings appear to be irrespective of geographical area (western countries vs. Asia) as well as head and neck tumor sites (oral cavity vs. oropharynx) and treatment approach (surgical vs. Radiation/chemoradiation) (13,35). Similar to previous studies, we found sarcopenia to be associated with the worse OS (HR 2.21 [95% CI 1.08 - 4.12], p = 0.03) in a cohort of 342 patients. In addition to the poorer prognosis of patients with sarcopenia, there is also increased risk of toxicity after treatment (10,19,35–37). Radiotherapy to the head and neck region is widely known to induce severe toxicities such as mucositis, odynophagia, and xerostomia, leading to critical weight loss and malnutrition (4,11,12,19). Although chemotherapy is not a primary treatment for HNC, it is often given in a concurrent setting. Recent retrospective studies in locally advanced HNSCC patients concluded that pre-treatment sarcopenia was a significant predictor of chemotherapy dose-limiting toxicity in patients treated with CRT using platinum-based chemotherapy (38,39). In this study, we tested the correlations between sarcopenia and a series of toxicity endpoints. We found sarcopenia was associated with longer PEG tube duration (median 162 days vs 134 days, HR 0.66 [95 %CI 0.48 – 0.89], p = 0.006) and higher risk of having PEG tube at last follow (OR 2.25 [95 %CI 1.12 - 2.08], p = 0.05). This agrees with the study by Karsten et al. that demonstrated sarcopenia contributes to the risk of prolonged feeding tube dependency of HNC patients treated with primary chemoradiotherapy (CRT) (37). Sarcopenia showed a non-significant trend towards higher risk of hospitalization < 3 months after RT (OR 2.18 [95% CI 0.82 - 5.79], *p* = 0.12). We did not see the association between sarcopenia with risk of osteoradionecrosis (p = 0.393) and post-RT stricture (*p* = 0.24). In HNC surgical populations, sarcopenia is a demonstrated negative prognostic indicator for both overall complications and wound complications and also pharyngocutaneous fistula in patients undergoing total laryngectomy for HNSCC (17). Yet in our study, sarcopenia was not associated with treatment-complication requiring surgery (*p* = 0.50).

Our study has several limitations that should be considered. Firstly, our analysis is limited by the inherent constraints of a retrospective study. Due to various exclusion criteria, such as missing pretreatment CT scans, missing clinical information, and scan artifacts, a large number of patients were excluded from our analysis, which may impact the distribution of patient characteristics. Secondly, our median dice scores were lower than those reported by Naser et al. (0.90 vs. 0.95) (27). We believe this is due to the preprocessing step we implemented to account for significant differences in CT imaging parameters, such as field of views, spacings, and slice thickness, between our development cohort (MDACC) and external test cohort (BWH). We were able to achieve a median DSC of 0.94 for validation and internal test sets in the MDACC cohort without this preprocessing step. However, the robustness of our model decreased when applied to the external dataset. Moving forward, we plan to optimize our imaging preprocessing steps to further improve our model’s performance while maintaining its generalizability. Moreover, we utilized the pre-defined gender-specific cut-off values proposed by Prado et al. (33) to determine sarcopenia. We found that female patients had significantly lower SMI values than males, consistent with previous studies. However, the cut-off values proposed by Prado et al. were based on an obese population (mean BMI 34.3 kg/m2), while the mean BMI in our population was 27.8 kg/m2. There is currently no consensus on the optimal method to define sarcopenia, and several other proposed thresholds exist. The end-to-end DL pipeline we developed for fully automated C3 segmentation allows for the efficient analysis of a large number of CT images from HNC patients. Traditional approaches involving manual or semi-automated C3 segmentation are laborious and require substantial expertise, making it challenging to analyze large datasets, particularly in multiinstitutional studies. In the future, we aim to expand our study to include international multi-institutional patient cohorts to identify optimal cut-off values for sarcopenia through further analyses, such as receiver-operating characteristics and precision-recall analyses. We hope this will establish a more reliable association between sarcopenia and clinical risk factors for HNSCC patients.

## CONCLUSIONS

We have developed and externally validated a fully-automated DL platform for fast and accurate sarcopenia assessment that can be used on routine head and neck CT imaging. Our model has demonstrated excellent C3 SM segmentation capability on datasets from different institutions, with high agreement with expert clinicians’ segmentation and high pass rates from expert clinicians’ reviews. Furthermore, we have shown that our model’s estimated SMI strongly correlates with the ground truth SMI. We have also demonstrated that SMIs predicted worse overall survival and longer PEG tube duration in a large HNSCC cohort from our institution. If further validated, our end-to-end DL pipeline has significant potential for incorporation into standard clinical practice for directing future treatment approaches and clinical decision-making, as well as for individualized supportive measures, including nutrition guidance or physical therapy.

## Data Availability

All data produced in the present study are available upon reasonable request to the authors

## ACKNOWLEDGEMENT

The authors acknowledge support from the National Institutes of Health (NIH) with grant numbers (NIH U24CA194354, NIH U01CA190234, NIH U01CA209414, NIH R35CA22052, NIH K08DE030216, NIH 5F31DE031502-02), the European Union-European Research Council (866504), and the Radiological Society of North America. All analyses and conclusions in this manuscript are the sole responsibility of the authors and do not necessarily reflect the opinions or views of the clinical trial investigators, the NCTN, or the NCI.

## AUTHOR CONTRIBUTIONS

Study design: B.H.K., Z.Y., A.S., Y.R.; code design, implementation and execution: Y.R., Z.Y.; acquisition, analysis or interpretation of data: Z.Y., A.S., B.H.K., Y.R., Y.Z., F.H., H.J.W.L.A., J.D.S., R.H.M.; image annotation: A.S., B.H.K., F.H.; writing of the manuscript: Z.Y., A.S., Y.R., B.H.K.; critical revision of the manuscript for important intellectual content: all authors; statistical analysis: Z.Y., Y.Z., A.S., Y.R.; study supervision: B.H.K., H.J.W.L.A., C.D.F.

## DATA AVAILABILITY

TCIA data including raw CT images may be requested from The Cancer Image Archive (https://www.cancerimagingarchive.net). Although raw CT imaging data cannot be shared, all measured results to replicate the statistical analysis will be shared once the manuscript is accepted for publication. Furthermore, we include test samples from a publicly available data set with deep learning and expert reader annotations.

## CODE AVAILABILITY

The code of the deep learning system, as well as the trained model and statistical analysis will be publicly available once the manuscript is accepted for publication.

