## supplement for "Fully-automated sarcopenia assessment in head and neck cancer: development and external validation of a deep learning pipeline" (from another version of this preprint: version 1, 2023-03-03)

### SUPPLEMENTARY MATERIALS

### SUPPLEMENTARY TABLES

**Table S1.** Summary on failing cases on external test set.

| Failing Causes | Failing Numbers (n = 23) |
|---|---|
| Missing sternocleidomastoid muscle (SCM) | 9 (39.1%) |
| Lymph node included | 6 (26.1%) |
| Post neck muscle missing | 3 (13%) |
| Anterior deep muscle missing | 3 (13%) |
| Submental muscle included | 1 (4.4%) |
| Other muscle included | 1 (4.4%) |

### SUPPLEMENTARY FIGURES

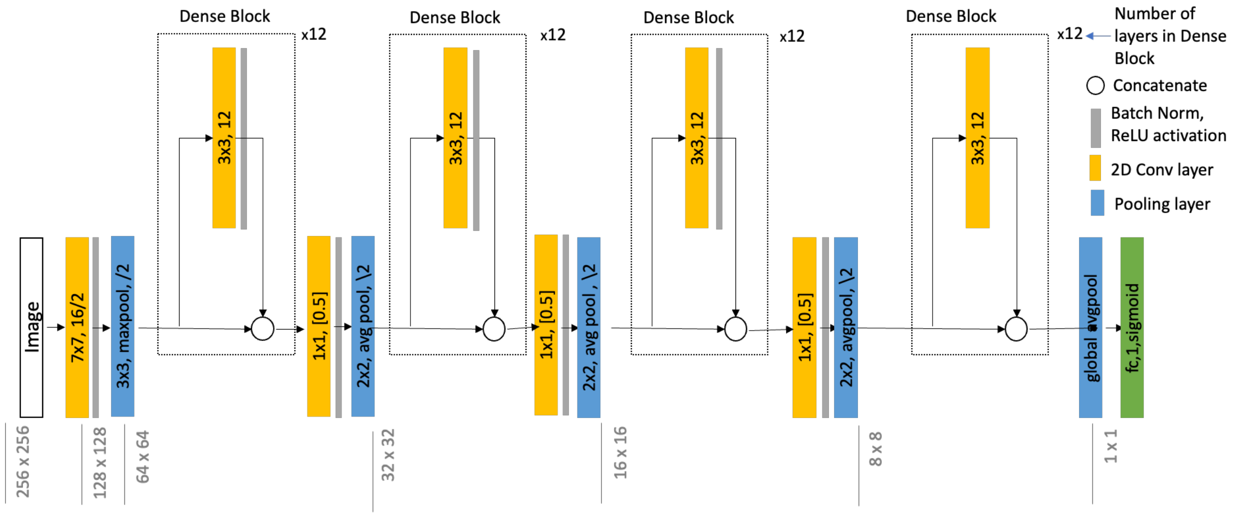


**Figure S1.** Model architecture for 2D DenseNet-based slice selection model. Adapted DenseNet CNN Architecture. Yellow blocks indicate 2D convolutional layers described by their kernel dimensions and number of output features. 'x12' indicates 12 layers within each Dense Block. Gray blocks indicate batch normalization layer followed by ReLu activation. Blue blocks represent pooling layers. '/2' indicates that the conv/pool layer has stride 2, otherwise the stride is 1. In the

DenseNet transition blocks '[0.5]' indicates the number of output features is half the number of input features (a compression factor of 0.5).

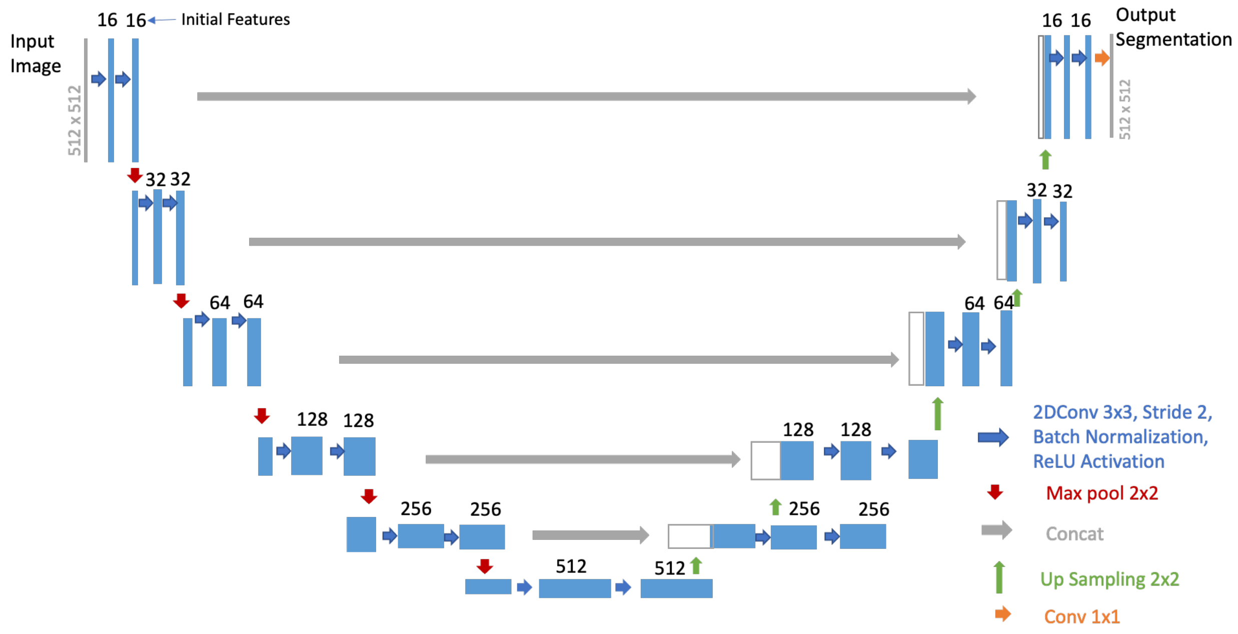


**Figure S2.** Model architecture for 2D U-Net-based segmentation model. The number of features specified above each block.

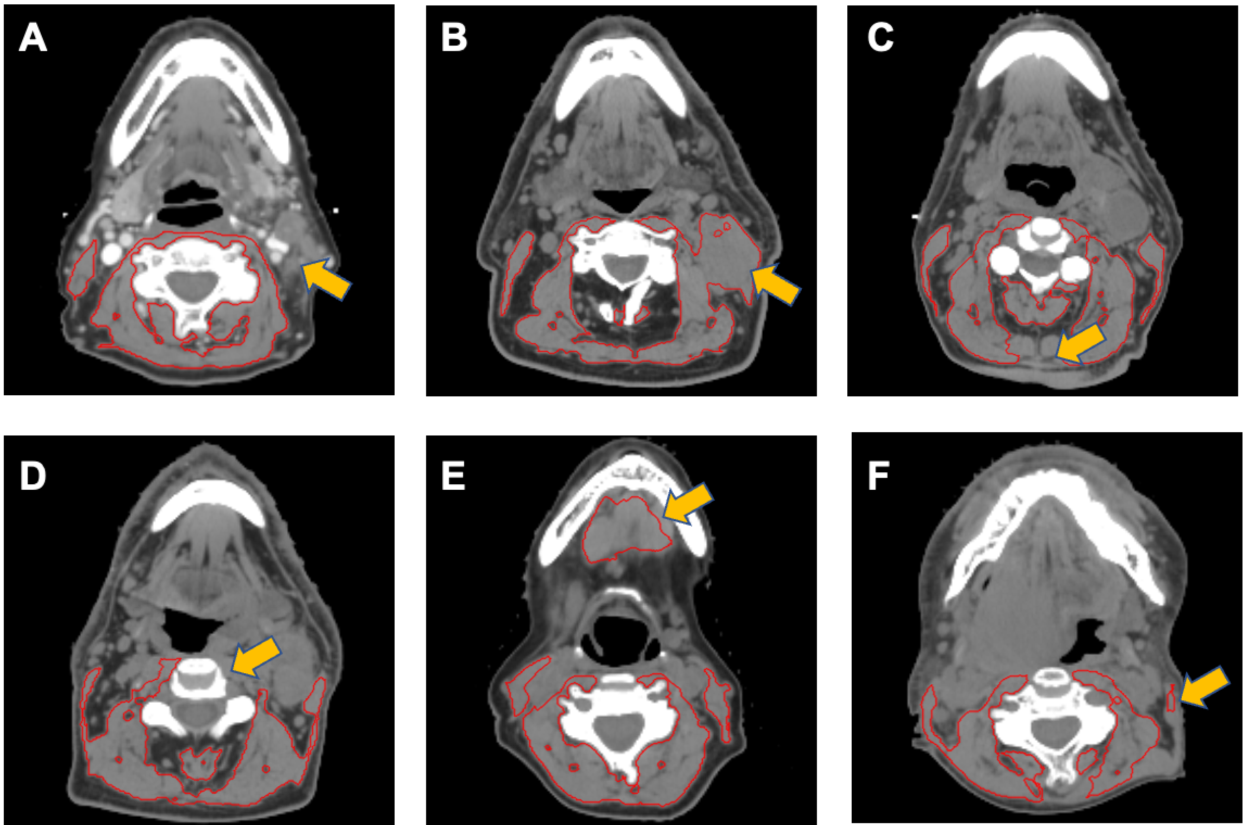


**Figure S3.** Representative failing cases with different failing causes. (A) Left sternocleidomastoid muscle (SCM) was missing; (B) Lymph node was included; (C) Post neck muscle was missing. (D) Anterior deep muscle was missing. (E) Submental muscle was included. (F) Other muscle was included.
